# Disease progression strikingly differs in research and real-world Parkinson’s populations

**DOI:** 10.1101/2024.02.17.24302981

**Authors:** Brett K. Beaulieu-Jones, Francesca Frau, Sylvie Bozzi, Karen J. Chandross, M Judith Peterschmitt, Caroline Cohen, Catherine Coulovrat, Dinesh Kumar, Mark J. Kruger, Scott L. Lipnick, Lane Fitzsimmons, Isaac S. Kohane, Clemens R. Scherzer

## Abstract

Characterization of Parkinson’s disease (PD) progression using real-world evidence could guide clinical trial design and identify subpopulations. Efforts to curate research populations, the increasing availability of real-world data and recent advances in natural language processing, particularly large language models, allow for a more granular comparison of populations and the methods of data collection describing these populations than previously possible.

This study includes two research populations and two real-world data derived (RWD) populations. The research populations are the Harvard Biomarkers Study (HBS, N = 935), a longitudinal biomarkers cohort study with in-person structured study visits; and Fox Insights (N = 36,660), an online self-survey-based research study of the Michael J. Fox Foundation. Real-world cohorts are the Optum Integrated Claims-electronic health records (N = 157,475), representing wide-scale linked medical and claims data and de-identified data from Mass General Brigham (MGB, N = 22,949), an academic hospital system. Structured, de-identified electronic health records data at MGB are supplemented using natural language processing with a large language model to extract measurements of PD progression. This extraction process is manually validated for accuracy.

Motor and cognitive progression scores change more rapidly in MGB than HBS (median survival until H&Y 3: 5.6 years vs. >10, p<0.001; mini-mental state exam median decline 0.28 vs. 0.11, p<0.001; and clinically recognized cognitive decline, p=0.001). In the real-world populations, patients are diagnosed more than eleven years later (RWD mean of 72.2 vs. research mean of 60.4, p<0.001). After diagnosis, in real-world cohorts, treatment with PD medications is initiated 2.3 years later on average (95% CI: [2.1-2.4]; p<0.001).

This study provides a detailed characterization of Parkinson’s progression in diverse populations. It delineates systemic divergences in the patient populations enrolled in research settings vs. patients in the real world. These divergences are likely due to a combination of selection bias and real population differences, but exact attribution of the causes is challenging using existing data. This study emphasizes a need to utilize multiple data sources and to diligently consider potential biases when planning, choosing data sources, and performing downstream tasks and analyses.

## Introduction

Despite substantial investment and increased biological understanding, disease-modifying therapies for Parkinson’s disease (PD) have not yet proven successful.^1^ Therapies recently considered to be promising in preclinical and early clinical trials have failed at later stages.^2^ There are numerous challenges to developing disease-modifying therapies for PD. Perhaps most importantly, we lack a complete understanding of the molecular processes driving disease pathogenesis.^3–5^ A wide range of PD onset ages and differing patterns of disease progression suggest heterogeneity.^6–8^ Recently, genome-wide survival studies have identified six loci --- *GBA*, *APOE*, *RIMS2*, *TMEM108*, *WWOX*, and the mitochondrial super macro haplogroup J,T,U --- associated with genetic heterogeneity in the progression from PD to Lewy body dementia.^9–13^ This is important because homogeneous patient populations and an accurate measurement of disease progression would greatly facilitate clinical trial design. More homogenous subpopulations may make biomarker associations more powerful, clarify the genetic basis for disease, and lead to more successful therapeutic development.^12,14^

Characterization of Parkinson’s disease (PD) progression using real-world evidence could guide clinical trial design and identify subpopulations. Most prior work examining PD progression focused on research data from major academic centers,^15^ but this may not accurately reflect the population at large.^8,16^ It is important to determine whether analyses performed in one particular data modality will lead to generalizable results. Large scale adoption of electronic health records and recent advances in natural language processing (NLP) could be used to address this knowledge gap. Individual health systems (e.g., Mass General Brigham, MGB)^17^ and large national managed care providers (e.g., UnitedHealth Group, Optum)^18^ have developed robust databases in the form of electronic health records (EHRs) through routine care. Even after filtering to enrich for newly diagnosed PD patients—those who had at least two years of data prior to diagnosis—Optum includes 157,475 patients with at least two PD diagnoses and MGB has a further 13,257 patients. It is important to note that while large, inclusion in these datasets is driven by access to care (e.g., insurance, socioeconomic factors) at MGB or within UnitedHealth Group. In addition to these new sources of real-world data (RWD), natural language processing and especially large language models have recently achieved improved performance on benchmark evaluations by substantial margins. In this work, we leverage these improved models to supplement structured EHR data at MGB by extracting phenotypic information, particularly clinical scores from clinical notes.^19–23^ To ensure the model’s extraction process is accurate, it is straightforward to perform manual chart review on a sample set of notes.

In this study we provide a comparison between two research studies and two RWD sources. While the research cohorts were actively recruited, the RWD are “passively” collected because they are derived from typical clinical care and not explicitly for research purposes. We characterized PD progression to better understand “typical” progression across the four research and real-world data sources. We evaluated the strengths and weaknesses of these data sources to characterize PD progression and their potential to enable more precise endophenotyping.

In performing this evaluation, we acknowledge and emphasize substantial potential sources of bias related to the way the different datasets were collected. For example, in the Fox Insight study, patients are recruited through online advertising, and they provide their data through an online portal. Within HBS, patients must have a Mini-Mental State Examination (MMSE) score consistent with the ability to provide informed consent or their next of kin must be present to provide consent. While ethically critical, criteria like this may systematically exclude certain participants, those that are older may be less likely to have and use the internet on their own, may have lower MMSE scores and be less likely to be in close contact or proximity with their next of kin. On the other hand, RWD are available only in the context of an individual’s interactions with healthcare and can be biased by access to healthcare and other socioeconomic factors. Additionally, analyzing RWD requires specifying rules for inclusion, clinical event, and outcome definitions. Reasonable changes to these rules have the potential to significantly change results.^24^

This study examines the differences of PD progression between these data modalities with the intent of providing guidance for future studies. By better understanding potential biases in data, future study of potential disease-modifying therapies can be designed to minimize these biases. This can be done by choosing the appropriate data for the question at hand and including multiple data modalities to help determine whether effects are artifacts of the data source or accurate measures of progression. This has the potential to aid future studies by providing guidance of when changes in progression are significant, the determination of appropriate clinical endpoints, and baselines for the identification of patient subpopulations of interest.

## Results

The study took place across four different datasets, two research populations: the 1) Harvard Biomarkers Study (HBS), and the 2) Fox Insight longitudinal health study (Fox Insight); and two real-world populations: 3) Mass General Brigham electronic medical records (MGB), and 4) Optum Claims-EHR dataset (Optum). This study was approved by the Mass General Brigham IRB. HBS, Fox Insight and Optum are de-identified and MGB electronic medical records were accessed through the Research Patient Data Registry. All work was retrospective review without additional contact or intervention in accordance with the IRB. No linking between datasets was performed.

The research populations (HBS and Fox Insight) had lower age of PD diagnosis than the real-world populations (mean 60.4 vs. 72.2, p < 0.001, Table 1). Fox Insight had a shorter length of follow-up after enrollment, limited by the fact that it is a new resource. HBS demonstrated the potential for research data sources to offer significant longitudinal data that is thus far difficult to find in RWD (mean follow-up time of 6.1 years vs. 3.2 years, p < 0.001). Finally, because the research cohorts were actively enrolled, there is the potential to collect additional history, for example the timing of an initial PD diagnosis is available for all participants of HBS and Fox Insight, but it must be extracted with rules from the RWD sources.

**Table 1.**
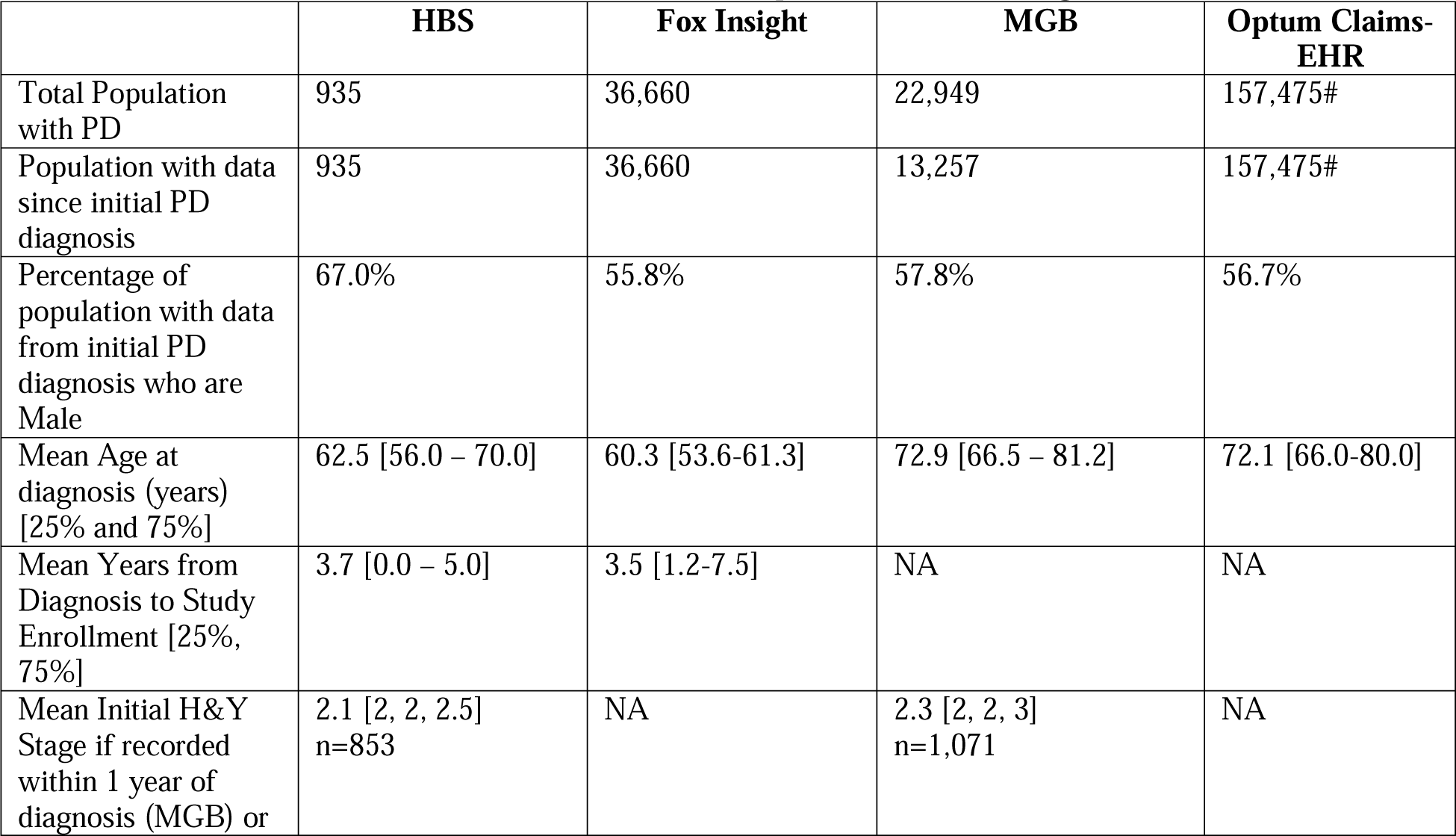
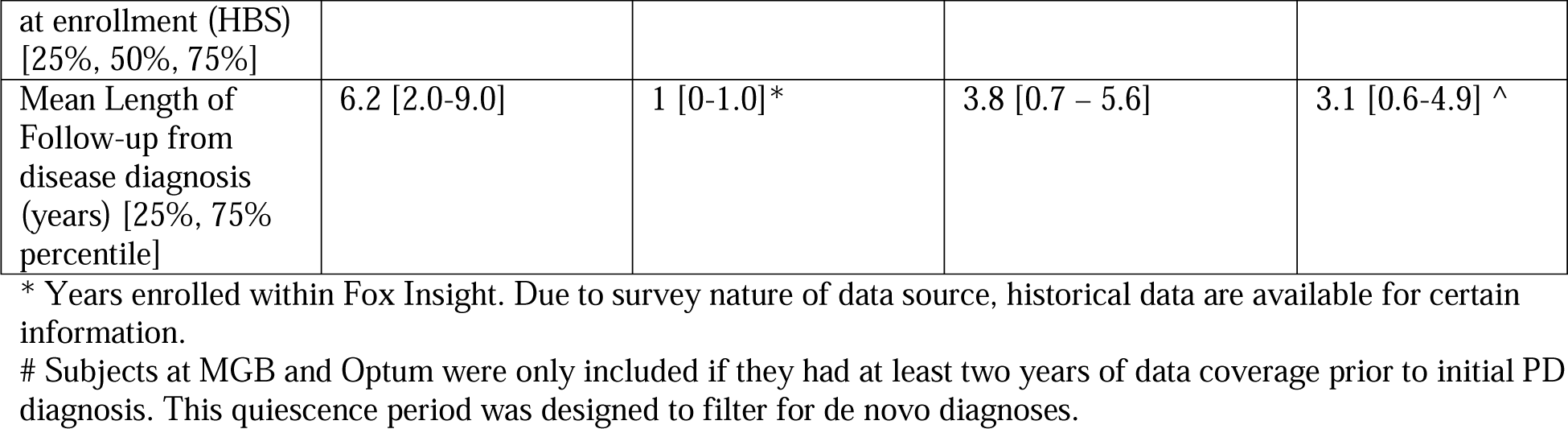
Cohort Characteristics for HBS, MGB, Optum, and Fox Insights cohorts.

Within HBS, clinical rating scales are actively collected, recorded as structured data and are available in most patients (Table 1, 853/935: 91.2%). In MGB they are passively collected and were extracted from the clinical notes for a small proportion of patients (1,071/13,257: 8.1%, extraction process described in methods). Clinical scores are not available in Optum. To evaluate the effects of potential interventions, it is important to understand the typical progression of PD. Clinical rating scales, including H&Y, MMSE, UPDRS and MoCA represent common clinical study endpoints for Parkinson’s progression. We compared progression of these measures between HBS, an actively assessed research cohort and MGB, a real-world cohort where data are available only if they are collected as a part of routine clinical care (i.e., passive data collection). We compared the cohorts by stratifying patients based on their time from initial diagnosis and compared clinical scores (Figure 1). We observe several key results: first, scores within HBS show slower movement than MGB in general than HBS. Outside of the UPDRS total score for MGB, the scores do not change substantially during the length of time of typical clinical trials (1-5 years). MMSE was significantly higher throughout the first five years within HBS (p<0.001, Figure 1C). Finally, the biggest change in progression is observed in the UPDRS total score, with the median score at MGB doubling by year 6.

**Figure 1.**
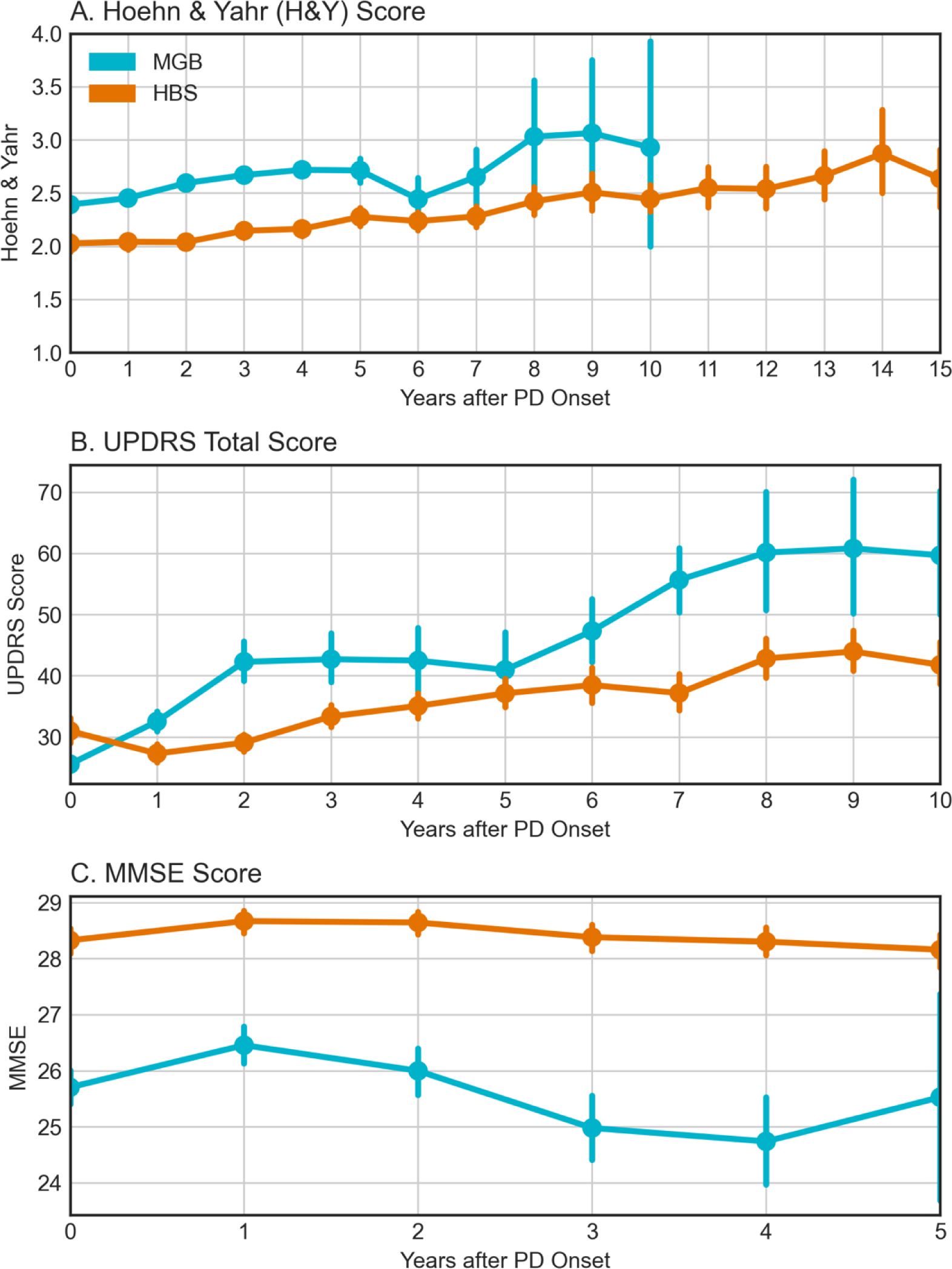
Parkinson’s Progression as measured by clinical rating scales (Teal = MGB, Orange = HBS). MGB were only available for up to ten years due to patient size drop-off. **A.)** Hoehn & Yahr Stage (MGB & HBS), **B.)** UPDRS Total Score (MGB & HBS), **C.)** MMSE Score (MGB & HBS),

Survival against key clinical events including levodopa initiation, reaching H&Y Stage 3, and discharge to long term care, first falls, or fractures after PD diagnosis, diagnoses of depression, and diagnoses of dyskinesias were evaluated in each of the data sources. We observed significantly earlier start of PD medications in the research populations compare to the real-world populations (p < 0.001). This was particularly pronounced in the Fox Insights data (Figure 2A). A potential explanation for this outlier result in Fox Insights could be that it entirely relies on patient self-reports. Interestingly, patients in the EMR data (MGB) were overall started on PD medications later than the research cohorts, however, initiation of levodopa therapy specifically overlapped with the research cohorts (Figure 2B). This may indicate that newer non-levodopa PD medications may be more aggressively prescribed in the patients participating in the research cohorts. Patients in RWD reached a H&Y score of 3 or more (Figure 2C, p < 0.001) or matched the definition for cognitive decline (Figure 2D, Optum vs. HBS, p < 0.001, MGB vs. HBS, p=0.001) sooner after diagnosis than those in the research settings.

**Figure 2.**
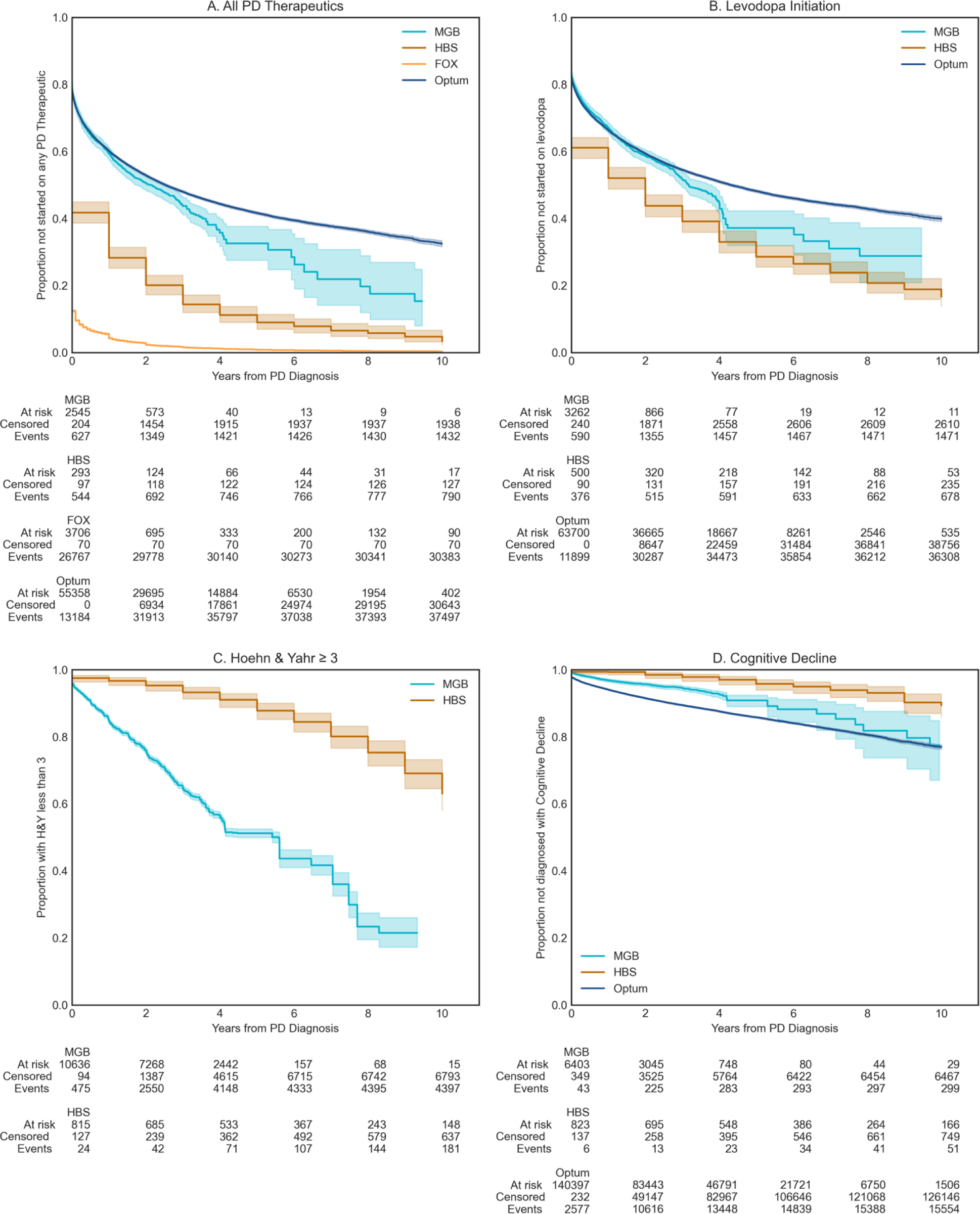
Survival against key clinical events. **A.)** Proportion of patients not started on Levodopa, **B.)** Proportion of patients not started on PD medications, **C.)** Proportion of patients with HY ≤ 3 (smaller or equal) (MGB, HBS), **D.)** Proportion of patients who have not experienced Cognitive Decline Proxy (MGB, HBS, Optum)

Additionally, we examined the association between disease duration (e.g., years since PD diagnosis) and the clinical rating scale score conditioning on the age of diagnosis, sex, and number of years of education (years of education is available in HBS only). For UPDRS, after adjusting for covariates, the intercept was 18.93 in MGB and 28.25 in HBS. At the intercept, the MGB slope was more than double that of HBS (3.87 vs. 1.54 within HBS, p < 0.001). Prior studies have estimated a 3 to 14 total UPDRS change per year, making the observed values in HBS lower than previously reported.^25^ Slopes for H&Y were not observed to be significantly different between MGB and HBS (Supplemental Table 4). While H&Y ranges from 1-5, few patients were observed on either extreme, indicating that this measure may not be sensitive enough to observe differences in the follow-up period. Finally, after adjusting for covariates, significant differences were observed between MMSE scores both at the time of diagnosis (HBS: 28.7, MGB: 25.8) and in terms of progression (HBS: 0.11 annual decline, MGB: 0.28 annual decline). Thus, both survival models and the linear regression model (Supplemental Table 4) are consistent with more rapid motor and cognitive decline in real-world vs. research population data. The change in H&Y is in line with what is previously reported in the literature (Supplemental Table 5).

Additionally, we broke down the percentage of patients who started therapy with levodopa by their H&Y stage (Supplemental Table 6). By H&Y stage 2.5 >80% of patients had initiated levodopa and we did not see any further increase. This is consistent with clinical guidelines indicating levodopa should be started in patients over 65 upon mild symptoms and for all symptoms as moderate or severe symptoms present.^26^ This plateau of ∼80-85% levodopa initiation during the study follow-up period indicates an upper limit on the utility of the surrogate measure of levodopa initiation to track PD progression. Levodopa provides a benchmark for the treatment of many PD symptoms which can be used as a tool to compare experimental disease modifying therapies or to plan future clinical trials.

We then examined the incidence of complications with major effects on patients’ quality of life, that is, fractures and falls; and depression.^27,28^ We observed a substantially higher and earlier incidence of fractures and falls as well as depression diagnoses in Optum compared to MGB data (p < 0.001, Figure 3A, 3B). The interpretation of this divergence is complex. The difference could be due to distinct care utilization; primary vs. tertiary point of care; general neurologist vs. movement disorders specialty care; or other systemic differences in the patient populations such as socio-economical divergence. This result indicates the need to examine results from RWD in the context of healthcare delivery.

**Figure 3.**
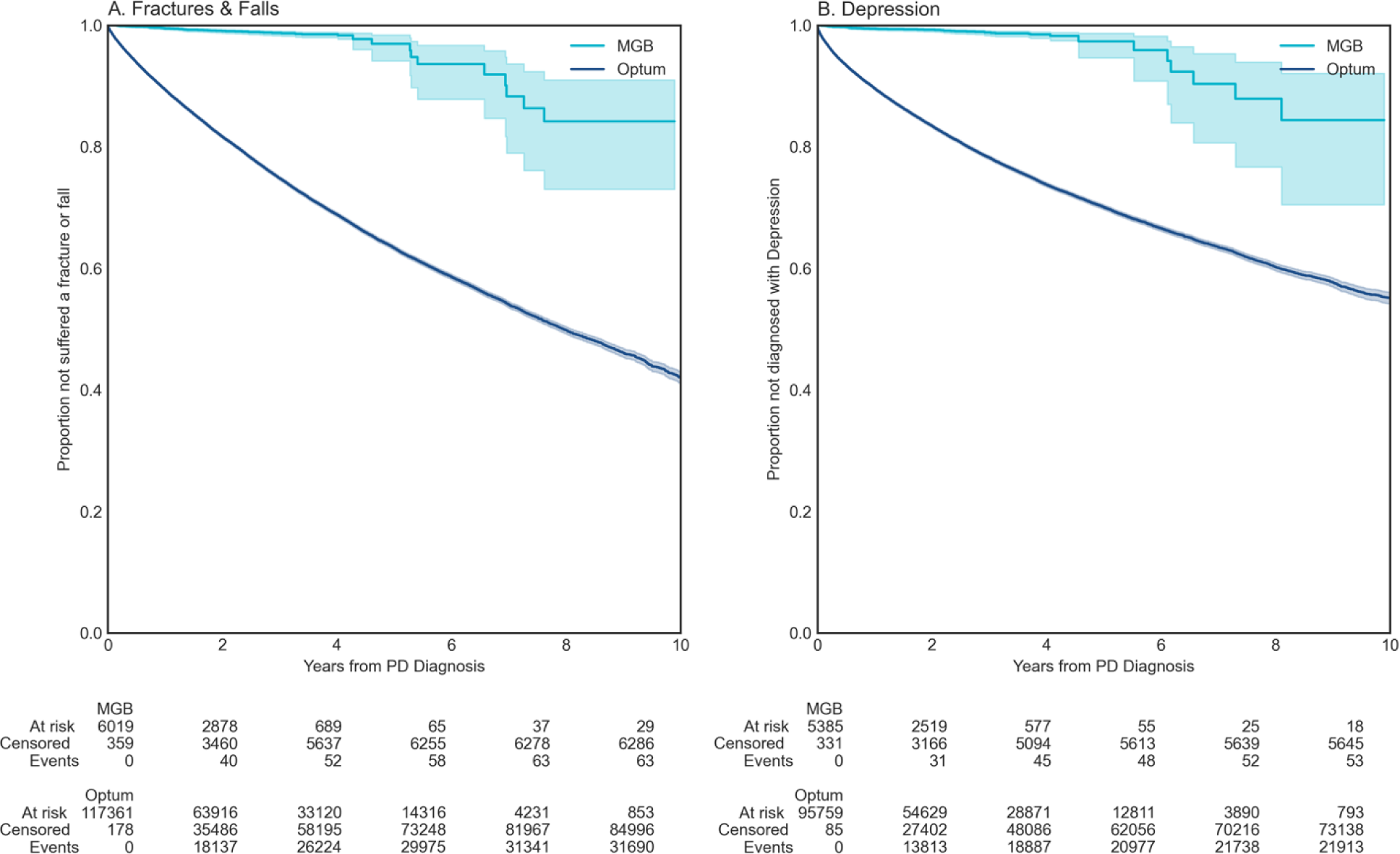
**Survival against indirect adverse events in RWD.** **A.)**, Fractures & Falls (MGB, Optum), **B.)** Depression (MGB, Optum).

## Discussion

This work examines PD progression across four different datasets, two research populations and two real-world data populations. It demonstrates the ability to utilize large language models to extract clinical rating scores from clinical notes as part of routine care and to compare these scores to a research-level, longitudinal clinical observational study. This work shows progression patterns in clinical rating scales and fit models to estimate “typical” progression across the irregularly spaced events common in healthcare. A key purpose of this study is to illustrate that while we observe these differences, it is difficult to attribute these differences to specific sources. They are likely to be driven at least in part by biases specific to each data source and collection mechanism, the populations within each data set as well as clinical care provided within each data source. By preforming these analyses across four different data modalities, this work aims to provide a diverse set of measures accessible to researchers with access to different data sources. We hope that it can help researchers choose the appropriate data modality, or ideally multiple modalities, for their research question and to approach their analyses paying careful attention to sources of bias.

The study shows systematic differences and potential directional biases between research and real-world data sets (Figure 4). Patients in research populations are diagnosed much earlier, are started earlier on levodopa and other PD medications, and have slower changes in clinical scales of motor and cognitive progression. RWD-based populations are diagnosed at an older age, start medications later than research cohorts and have faster changes in clinical scales and events geared at measuring motor and cognitive decline. RWD-based populations offer substantially larger sample sizes, do not require active recruitment, and may be available immediately for past populations. Most importantly, RWD-based populations may be more representative of routine clinical care and thus may be more reflective of the general population.

**Figure 4.**
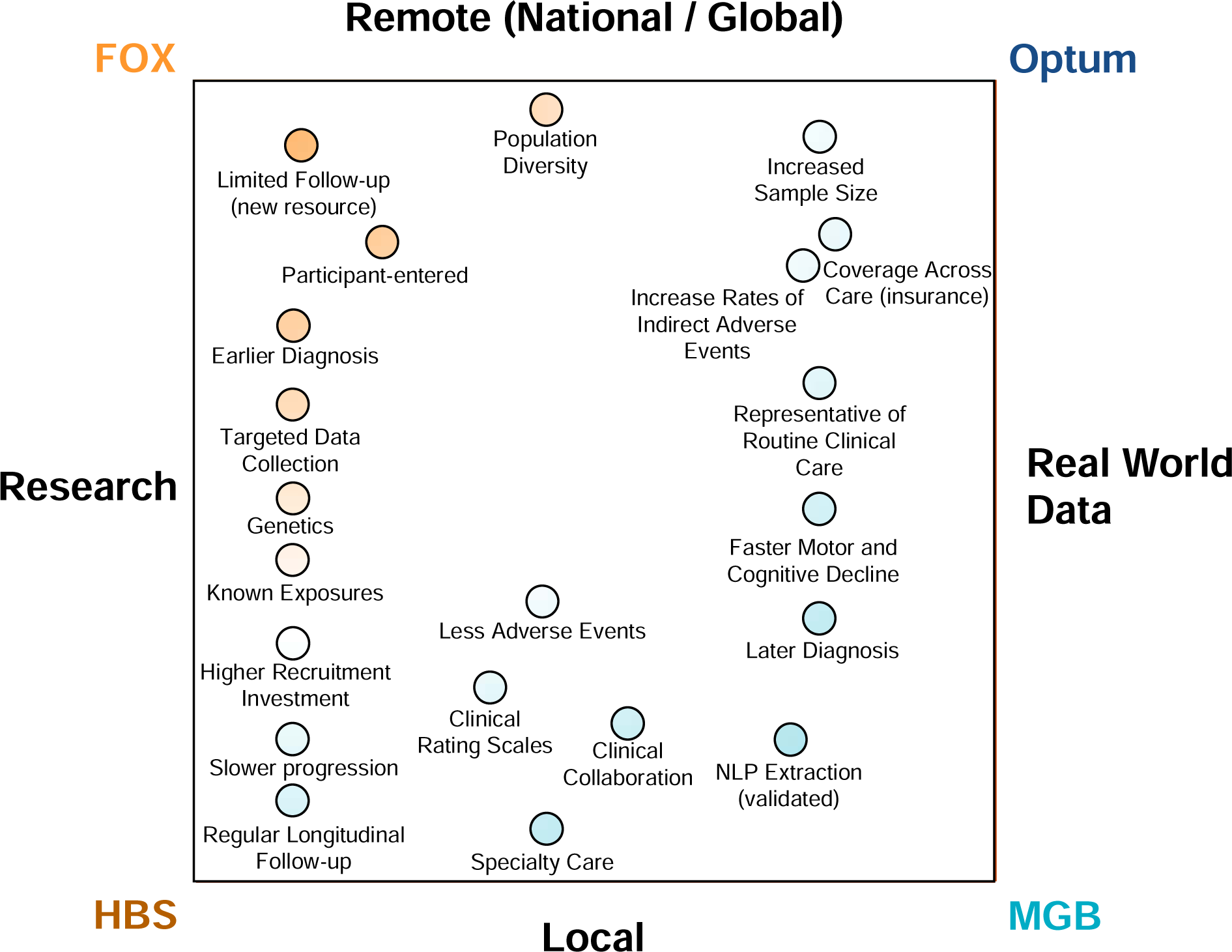
Comparison of PD phenotypic resources, research populations vs. real-world data and local vs. remote demonstrating the pros and cons of each data modality for PD research.

Which variables may underlie the divergent progression features in the real vs. the research world? We hypothesize that several factors may play a role:

1. The way cohorts were recruited (e.g., actively recruited research datasets or passively generated real world data extracted from routine clinical care). This may lead to recruitment bias for younger and more educated patients into research studies.^16,29^ We previously observed “proxy” diagnoses where a non-specialist might diagnose a patient with a less specific gait, tremor, or cognitive disorder as justification for a referral to a movement disorders specialist.^30^ One potential explanation for this pattern could be a hesitation to make a diagnosis as severe as PD without being completely confident. Populations enriched for Mendelian forms of PD have been observed to have an earlier disease onset and could be over-represented among the research populations. Their earlier diagnosis of PD may be the result of greater access to specialists resulting in a faster transition from a “proxy” diagnosis to PD. Due to critical ethical requirements, HBS requires cognitive competency or a next of kin to provide informed consent. This requirement may exclude clinical subtypes of PD more likely to have substantial cognitive decline before they are able to be recruited and enrolled.^31^ In contrast, patients with later diagnosis and patients with the postural instability-gait difficult (PIGD) form of PD have both been observed to have faster UPDRS part II^32^ as well as general cognitive^33^. There is a relationship between PIGD and disease duration^34,35^, so accurate assessment of disease onset is critical for future studies of subtypes. This study emphasizes the challenges in differentiating potential phenotypic subtypes from confounding factors such as disease duration.
2. The specialization of care, particularly in terms of what is deemed relevant for treating (MGB) or studying PD (HBS) may dictate which data are collected and how they are collected in comparison to networks that should have comprehensive coverage of all care (Optum). This discrepancy may be a large component of the differences in survival against depression as well as falls and fractures.
3. The source of the data (e.g., provider vs. patient provided) may have led to exaggerated differences between Fox Insight and the other data sources. This was especially relevant regarding PD-therapeutic initiation. In contrast to data from each of the other sources, most patients within Fox Insight believed they immediately started a PD-therapeutic. The different data sources also dictated which analyses were possible, this was especially seen in longitudinal analyses reliant on follow-up periods. Fox Insight represents a new data source and therefore the length of follow up was limited. However, HBS demonstrates the potential for research data sources to offer significant longitudinal data that is difficult to find in RWD (mean follow-up time of 6.1 years vs. 3.2 years, p < 0.001). This comparison between data modalities may be related and may help to quantify potential “hawthorne effects”, where greater interaction between participant and physician can lead to differing outcomes.^36,37^

As RWD becomes increasingly available, it is critical to understand the strengths and weaknesses of different sources of data and analytical approaches. This study identified important limitations for each of the datasets. HBS has strengths in terms of the quality of targeted data (e.g., environmental exposures, clinical rating scales, genetics) but given its nature as longitudinal biomarkers study with structured study visits, it is comparatively small in participant numbers. As it is based at a single site in Boston, it also may not be representative of the diversity of PD across the nation. MGB shares this issue of representation but has approximately 25 times as many participants as HBS (22,949 vs. 935) and includes data extracted from typical clinical care records for PD (i.e., real-world data). Patients at MGB and HBS are predominantly of European descent, while ethnicity data were not available in the licensed Optum dataset. This meant, that despite being large data sources, we were not able to subset people with PD according to race or ethnicity in search of different progression patterns. We do however, suspect that there are substantial biases related to access to care within both MGB and Optum. MGB is a highly ranked academic health system and those with means such as insurance, ability to travel, and the support of caregivers who can afford time away to help may be more likely to seek out care at MGB as opposed to whichever provider is closest. Similarly, while geographically broad, Optum is still highly enriched for private insurance and the Midwest region. These factors are not easy for single organizations, even organizations with as many resources as these to mitigate. Less biased data may require system-wide intervention or efforts such as government supported registries universal access to care. Studying neurodegenerative conditions that frequently emerge around the age of retirement is especially challenging as people frequently switch insurers or even move to new locations for retirement. In both cases, neither the claims data nor provider data alone can be relied upon. The population that has continuous data is likely to be biased by those that retire later and among other potential biases may be healthier or have increased socioeconomic needs to continue working.

Efforts such as Fox Insight have the potential to alleviate some of these challenges by recruiting for a large, diverse, patient population but this will require intentional effort. Importantly, Fox Insight also collects genetic data allowing for more accurate determination of ethnicity. Otherwise, limitations of EMR data sources like MGB include the fact that genetics are only available on a small subset of patients. Consequently, even where ethnicity data are available, analyses are often dependent on patient reported ethnicity rather than genetic ancestry. However, even this effort is subject to substantial bias since patients must interact with the study online, meaning the patient or someone close to the patient needs to have internet access. Adding to the access question, Fox Insight leverages online advertising and the populations who spend more time online may be more likely to discover Fox Insight.

Additionally, clinical scores are not typically captured in structured EHR data, there is no guarantee of completeness of care, there are censoring challenges, and these scores are based on subjective assessments. Future assessments using biomarkers (e.g., RT-QuIC^38^) may enable the measurement of progression in settings without experts trained in different clinical scales and may provide faster and more objective measurement of progression even when experts are available. Optum, as a combination of EMR and administrative claims data, improves upon the challenges of completeness of care data but has less per-patient granularity than identified EMR data. For example, in some cases it may be possible to extract clinical rating scores, but because end users do not have access to Optum’s proprietary natural language processing extraction code or the raw notes the values are extracted from, it’s not possible for an end user to validate the extraction. Within MGB it was possible to compare the extracted values to manual chart review-extracted values to validate that the NLP extraction process worked accurately. We are actively working to generalize this approach to share it as open-source software after evaluating in additional settings and cohorts. Without being able to validate the extraction process in Optum, it’s difficult to assess whether the extracted values can be used for downstream analyses with sufficient confidence. For this reason, we excluded the clinical rating scales available within Optum from our analyses. Despite these challenges, Optum includes nearly eight times the number of patients as a single site EMR like MGB and is likely more representative of national demographics of PD. We believe that future studies would benefit greatly from comparing both between RWD-sources as well as between the cohort of interest and background control populations. A limitation of this study was the lack of access to control populations in resources like Optum due to data licensing.

Finally, the Fox Insight study, is relatively new and therefore has limited follow-up time. Fox insight’s web-based recruitment and information collection model have demonstrated promise, rapidly recruiting tens of thousands of PD patients. Because it is survey-based, there is likely to be heterogeneity in response quality (e.g., therapeutic initiation), and unlike the other data sources, it is patient-provided. Data are not entered by physicians and clinical research affiliates. Additionally, Fox Insight, currently collects a history of the presence of clinical events but does not currently collect the date of occurrence. This may be difficult for subjects to remember but would be useful in characterizing progression (i.e., survival analyses require dates of clinical events). We hope that this work can provide guidance to future efforts of this emerging resource as well as aid in the design of future studies.

Due to the heterogeneity of PD, a better understanding of the “ground truth” of progression is critical for the development of disease-modifying therapeutics. This understanding may enable the identification of subpopulations for target discovery and improve the design of clinical trials (e.g., selection of more appropriate endpoints). Finally, without a better understanding of current progression, it is difficult to value therapeutics which may slow progression according to patient quality of life and healthcare cost savings.

## Methods

### Data Sources and Study Population

#### Harvard Biomarkers Study

The Harvard Biomarkers Study (HBS) is a longitudinal case-control registry which aims to accelerate the development of molecular diagnostics which track or predict progression of early-stage PD.^10,39–41^ The study includes participants with PD, mild cognitive impairment, Alzheimer’s disease in addition to healthy controls. Participants are recruited from throughout Mass General Brigham and enroll for at least three years though the Memory and Movement Disorders Units at Brigham & Women’s Hospital and Massachusetts General Hospital. Full study information is available in study publications^9,40,42^ and online (https://www.bwhparkinsoncenter.org/).

PD cases are defined according to the UK PD Society Brain Bank Criteria^1^ or a movement disorders specialists’ assessment. Cases were required to have Mini-Mental State Examination (MMSE) scores >21 or the next of kin present to provide informed consent. PD cases were excluded for presence of blood disorders, hematocrit levels <30%, and active ulcers or colitis. Clinical disease severity is measured in the “operational on state” according to clinician expert judgement, but patients were not necessarily at the peak response (“true on state”).

At the time of this analysis, the study included a total of 1,914 participants including 935 with PD and 979 healthy controls. The biobank includes genotyping (e.g., 42 with *GBA* mutations), medical record data, as well as repeat clinical score and survey measurements. Within HBS, year of diagnosis (but not day and month) were recorded. Thus, all analyses are done at the year of diagnosis level.

#### Fox Insight

Fox Insight is a web questionnaire-based, longitudinal study including both people having and not having Parkinson’s disease (see publication for full description)^43^. At the time of study, the Fox Insight database included 36,660 people with Parkinson’s disease. The questionnaire covers the patient’s medical and family histories, potential environmental exposures, as well as ongoing functional status. Additionally, genetic testing has been performed on a subset of the population.

#### Mass General Brigham (MGB) Electronic Medical Record

The MGB EMR (previously known as Partner’s Health Care) includes > 6.8 million total active records from 1990 to 2020. Available data include encounters, diagnoses, medications, procedures, observations, and labs. The MGB EMR also includes a biobank with >36,000 patients who have been genotyped with Illumina Multi-Ethnic Global Array (MEGA) microarray chip.

The full EMR includes 22,949 patients with at least two diagnoses of PD as in Yuan et al.^30^ (ICD9: 332, 332.0, ICD10: G20). To identify new onset PD, we required a two-year quiescence period (730 days) where a patient had encounters with the health system prior to having an initial diagnosis of PD and consider the time from initial (Supplemental Figure 1). There were 13,257 with encounters at least two years prior to their initial PD diagnosis. Full criteria for initial PD diagnosis are in the Study Design and Statistical Analysis section under *Initial Parkinson’s Disease Diagnosis*.

#### Optum® Integrated Claims-EHR Data

Optum-provided description (quoted directly)^44^: “Optum’s Integrated Claims-Clinical dataset combines adjudicated claims data with Optum’s Electronic Health Record data. Optum’s longitudinal clinical and claims repository from Optum Analytics is derived from more than 50 healthcare provider organizations in the United States, that include more than 700 Hospitals and 7000 Clinics: treating more than 102 million patients receiving care in the United States. Optum’s Integrated Claims-EHR dataset is statistically de-identified under the Expert Determination method consistent with HIPAA and managed according to Optum® and customer data use agreements. The Integrated dataset, which leverages a proprietary Optum algorithm that uses both salting and cryptographic hashing, links both claims and clinical data for approximately 24M matched individuals. The Integrated dataset includes historical, linked administrative claim data from pharmacy claims, physician claims, and facility claims, with EHR information, inclusive of medications prescribed and administered, lab results, vital signs, body measurements, diagnoses, procedures, and information derived from clinical notes using Natural Language Processing.”

We identified a total of 157,475 patients with at least two distinct PD diagnoses after at least 730 days between coverage beginning and an initial PD diagnosis. Available data for this cohort include encounters, diagnoses, medications procedures, observations, and labs. Full criteria for initial PD diagnosis are in the Study Design and Statistical Analysis section under *Initial Parkinson’s Disease Diagnosis*.

### Study Design and Statistical Analysis

#### Natural Language Processing Extraction of Clinical Rating Scales

We evaluated PD progression in accordance with common clinical scales: Hoehn & Yahr (H&Y)^45^, Mini–Mental State Examination (MMSE)^46^, Montreal Cognitive Assessment (MoCA)^47^, and the Unified Parkinson’s Disease Rating Scale (UPDRS).^48^ Scores were available as structured data within HBS but needed to be extracted at MGB using natural language processing. While each scale includes subjective assessments, they have undergone validation studies^10,48–58^ and are commonly used in the assessment of PD and/or general cognitive progression.

To extract scores, we used the Flan-PaLM instruction-finetuned model^22^ (flan-t5-xxl) as the base for a few shot (n=10 with clinical scores, 10 without clinical scores) approach with exemplars. The prompt provided exemplars of a positive example and a negative example with the form (italics replaced with actual note content):

“Q: Answer the following question with a value between 1 and 5 or NA. What is the Hoehn & Yahr score for the patient in the following note? Note: *Note Content* A: *3* Q: Answer the following question with a value between 1 and 5 or NA. What is the Hoehn & Yahr score for the patient in the following note? Note: *Note Content* A:”

We provided 10 prompts with exemplars that had clinical rating values present and 10 prompts with exemplars that did not have clinical rating scales for each extraction. We then compared the outputs between the inferences and flagged any notes with less than 9 out of 10 matching outputs. To verify that the model was appropriately finding scores when they were present, we manually reviewed the extracted scores for:

Hoehn & Yahr: 200 notes where the reviewer identified a score, 200 notes where the model identified a score, 200 notes where no score was found by the model and 200 notes where we manually identified that no score was found. For the 800 notes, extraction accuracy was 99.75%; 2 notes were incorrect. Both cases were appropriately flagged, as multiple Hoehn & Yahr values were recorded, and across the 10 runs only 4/10 and 5/10 scores matched.

UPDRS: 100 notes where the reviewer identified a score, 100 notes where the model identified a score, 100 notes where no score was found by the model and 100 notes where we manually identified that no score was found. For the 400 notes, model accuracy was 98%.

MMSE: 100 notes where the reviewer identified a score, 100 notes where the model identified a score, 100 notes where no score was found by the model and 100 notes where we manually identified that no score was found. For the 400 notes model accuracy was 99.50%

Across all 3 scores, no extraction with 9 or more matching outputs was incorrect in the randomly sampled evaluation sets. We therefore focused on reviewing all notes not included in our manual review pool that had inconsistent outputs across exemplar runs. This included 133 H&Y, 41 UPDRS, and 37 MMSE notes. The majority prediction (but less than 9) was correct for 94/133 H&Y notes, 36/41 UPDRS notes and 29/37 MMSE notes. We manually corrected the notes that did not meet our threshold with the human annotated value.

### Definitions of Health Care Utilization and Clinical Events

Other definitions used to describe healthcare utilization and clinical events are provided in full in Supplemental Tables 1-3 and described here.

#### Initial Parkinson’s Disease Diagnosis

The initial Parkinson’s disease diagnosis date is explicitly stated in both of the research data cohorts (HBS, Fox Insight). Within the RWD, we required a 730-day quiescence period to enrich for newly diagnosed PD (Supplemental Figure 1). Within Optum, this quiescence period was defined by taking the first observed PD diagnosis (Defined in Supplemental Table 2) and checking to see whether the subject had continuous coverage for the 730 days prior. To be included within MGB, an individual needed to have at least one encounter 730 or more days prior. In both RWD sources, subjects were then excluded if they did not have an additional PD diagnosis at least 30 days after the initial PD diagnosis in an attempt to filter out codes used for referral or diagnostic purposes.

Within the RWD, subjects diagnosed with conditions that commonly result in clinical parkinsonism symptoms during the two-year quiescence period were excluded.^30^ This included those with encephalitis, Alzheimer’s disease, or other cognitive conditions mimicking idiopathic PD. Additionally those with conditions such as schizophrenia, Parkinson’s-related disorders, metabolic neurogenic anomalies (e.g., Wilson’s Disease), or other degenerative ailments resulting in clinical parkinsonism symptoms (e.g., Multiple system atrophy, Progressive supranuclear palsy) were also excluded. The codes corresponding to these conditions are detailed in Supplementary Table 2 and derived from Yuan et al.^30^

This exclusion was not performed within HBS as subjects were screened by study personnel and it was not possible to perform within Fox Insights.

#### Cognitive Decline

Cognitive decline was defined as any diagnosis of mild cognitive impairment (ICD9: 331.83, ICD10: G31.84) a prescription for a medication typically used to treat cognitive impairment during an encounter with a PD diagnosis (Aricept, Exelon, Namenda, Galantamine using national drug codes derived with RxNorm^59,60^), or a referral for a neurology specialist for cognitive decline. For HBS, an MMSE score with the cut-off of ≤ 25 was defined as an indicator of significant global cognitive impairment as recommended by the International Parkinson and Movement Disorders Society (MDS) Task Force. The RWD definition was developed in conjunction with clinicians (led by CRS) and trial statisticians (JP, CC) to correlate as closely as possible to the definition using MMSE with data available in RWD. To validate this metric, we employed the Phi coefficient (L), to quantify the degree of association between two binary variables (matching cognitive decline definition, and MMSE threshold). We compared this coefficient for 200 patients across 912 visits where both the RWD-cognitive decline definition was available and a MMSE score is available. We obtained a L value of 0.848, suggesting a strong correlation between the two definitions.

#### PD Therapeutics & Levodopa

Within Fox Insights and HBS the timing of PD therapeutic is explicitly asked of the participant or entered by study staff respectively. Within HBS Levodopa initiation is explicitly asked, but within Fox Insights it is not. Therefore, Fox Insights was not included in the analyses for levodopa initiation.

For the RWD (Optum, MGB), PD therapeutic initiation (Supplemental Table 1) and levodopa initiation was considered to be first date of a sequence prescription fills meeting all of the following criteria:

a. Initial fill of a therapeutic listed in linked to an encounter with a PD diagnosis.
b. > 90 total days of pills.
c. 2+ fills occurring over a period of > 90 days but within 180 days from the first fill of the sequence.

These criteria aim to a.) ensure that the prescription is for PD, b.) the patient is not receiving the medication as part of an evaluation or receiving a single 90-day fill, and c.) identify the true starting point of treatment as opposed to evaluation.

#### Clinical Events - Depression & Falls and Fractures

Adverse clinical events were represented by operational criteria for health care utilization and are defined in Supplemental Table 3. These criteria were developed by the Health Value & Economics team within Sanofi.^61^ Patients were required to have at least one code for the specified event as either the primary or admitting code for a visit which included an inpatient stay or treatment (procedures or medications). Patients were excluded from this analysis if they experienced diagnoses for the clinical event in question during the quiescence period.

### Statistical Analyses

#### Statistical Tests and Survival Analyses

Quantitative population attributes (e.g., age of diagnosis) were first plotted using Quantile-Quantile Plots followed by a Shapiro-Wilk test for normality. After determining normality, Levene’s test was used to assess whether variances were equal. In the case of age and follow-up time, the variances were not equal and therefore a Welch’s t-test was performed to determine whether there were significant differences between the populations.

For Clinical rating scores (MMSE), after determining that the values were not normally distributed, a Mann-Whitney U test was employed to compare scores between HBS and the extracted scores from MGB.

Survival analyses against clinical events were conducted using the Kaplan-Meier method to estimate the time-to-event distributions. These analyses were performed using the ‘lifelines’ python package.^62^ For each patient, the time to clinical event (or last observed encounter) was recorded along with an binary indicator denoting whether the event was observed or censored. The resulting Kaplan-Meier survival curves visualize the time-to-event distributions, and we used a log-rank test to compare these distributions between different groups.

Finally, to ascertain an average progression of UPDRS, MMSE, and H&Y we fit a linear regression model accounting for the irregularity of sampling intervals. To do this we included the age at diagnosis, time since diagnosis, sex, and number of years of education (within HBS only) as covariates and fit a model to the appropriate clinical rating scale. Residual plots were examined to verify the homoscedasticity and linearity assumptions. The normality of residuals was assessed using quantile-quantile (QQ) plots and the Shapiro-Wilk test. Multicollinearity among predictors was evaluated using the variance inflation factor (VIF), ensuring all VIF values were below 10.

#### Missing Data

Within the two research databases, there was minimal missing data due to the overlap of their study designs and the analyses in this work. Within HBS, data are entered by study personnel who are instructed to complete all of fields used in this study. All participants had an age at study visit, age at diagnosis, gender, race, and ethnicity at each study visit. Across all study visits, H&Y was recorded in all visits and MMSE was recorded in all but 8 visits (99.8%). Visits without a MMSE value were excluded from analyses. Within Fox Insights, participants answer questions in panels in order as they sign up. Participants were excluded if they did not complete the initial set of questions required for this study. In the RWD, data are only available through normal clinical care when patients have interactions with the healthcare system. Study inclusion criteria were selected to maximize the likelihood that patient encounters would be captured. To this end, patients were required to either have coverage (Optum) or have prior visits (MGB) at least two years prior to their initial PD diagnosis. This was put in place to both filter for new diagnoses of PD as well as to limit patients who were referred after an initial diagnosis.

Additionally, many visits do not include a full PD-specific work up and/or do not record PD clinical rating scores. Less than 10% of patients had an initial H&Y score included in their note (1,071 of 13,257 patients, 8.08%). Similar to prior work^63^, we trained simple machine learning classifiers to predict whether a score would be present based on a patient’s age, sex, age at initial diagnosis, binary presence of comorbidities (rolled up to PheWAS codes),^64^ distance from the clinic, which clinic, and physician specialty. Outside of physician specialty, no feature was determined to be significant and even with physician specialty, performance was only marginally better than random (AUC: 0.56). Due to the relative rarity, lack of a clear discernable pattern of presence, and the fact that it could not be determined that missingness was dependent on outcome, we elected not to perform imputation and instead to include clinical rating scores only where they were available in the notes. This approach comes with an important caveat, the patients with scores may not be representative of the entire cohort. The reasons for missing scores may not be observed, whether related to disease severity, accessibility to care, or other clinical factors, could systematically differ between those with and without scores. This study aims to mitigate these challenges by using multiple datasets and presents a comparison between these datasets to better understand these biases.

### Standard Protocol Approvals, Registrations, and Patient Consents

This study was approved by the Mass General Brigham Institutional Review Board (#2017P002452). This study received only de-identified, retrospective data from research datasets (HBS and Fox). Research dataset participants are consented for study and publication as part of the recruitment process. Real world datasets (MGB and Optum) are retrospective de-identified data and therefore individual consent is not required for publication.

## Data Availability

Authors had primary access to all data sources used in this study (BKB). All data used in this study are secondary use and therefore the authors must direct those interested in using any data involved to their source:

1. Harvard Biomarkers set is accessible through Accelerating Medicines Partnership program for Parkinson’s disease https://amp-pd.org/unified-cohorts/hbs and through https://www.bwhparkinsoncenter.org/biobank/
2. Mass General Brigham EMR; https://rc.partners.org/about/who-we-are-risc/research-patient-data-registry
3. Optum® Integrated Claims-EHR Data^44^ (https://www.optum.com/content/dam/optum3/optum/en/resources/fact-sheets/Integrated-Data-product-sheet.pdf),
4. Fox Insight^43^ https://foxinsight.michaeljfox.org/

## Author Contributions

|  | Conception<br>or design<br>of the work | Acquisition<br>of data | Analysis<br>of Data | Interpretation<br>of Data | Manuscript<br>draft | Revision |
| --- | --- | --- | --- | --- | --- | --- |
| Brett K.<br>Beaulieu-Jones<sup>1</sup> | x | x (pre-<br>processing) | x | x | x | x |
| Francesca<br>Frau | x |  | x | x |  | x |
| Sylvie Bozzi | x |  | x | x | x | x |
| Karen J.<br>Chandross | x |  |  | x | x | x |
| M Judith<br>Peterschmitt |  |  | x | x | x | x |
| Caroline<br>Cohen |  |  | x | x | x | x |
| Catherine<br>Coulovrat |  |  | x | x | x | x |
| Dinesh<br>Kumar |  |  |  | x |  | x |
| Mark<br>Kruger |  | x |  | x | x | x |
| Scott L.<br>Lipnick | x | x |  |  | x | x |
| Lane<br>Fitzsimmons |  |  |  |  | x | x |
| Isaac S.<br>Kohane | x |  |  | x | x | x |
| Clemens R.<br>Scherzer | x | x |  | x | x | x |

## Acknowledgements

This work was funded in part by the Sanofi iDEA Awards Initiative (BKB) as well the National Institute of Neurological Disorders and Stroke grant number (K99NS114850, R00NS114850). CRS’s work is supported in part by NIH grants NINDS/NIA R01NS115144, U01NS095736, U01NS100603, the U.S. Department of Defense, and the American Parkinson Disease Association Center for Advanced Parkinson Research. The Fox Insight Study (FI) is funded by The Michael J. Fox Foundation for Parkinson’s Research. This research was funded in part by Aligning Science Across Parkinson’s [ASAP-000301] through the Michael J. Fox Foundation for Parkinson’s Research (MJFF). We would like to thank the Parkinson’s community for participating in this study to make this research possible. We would like to acknowledge Cliona Molony and Sanofi Digital, for facilitating data access through the Sanofi Darwin platform to support this project. We would also like to thank all of the participants and organizers of the Harvard Biomarker Study as well as the organizers of the Research Patient Data Registry at Mass General Brigham. For the purpose of open access, the author has applied a CC BY public copyright license to all Author Accepted Manuscripts arising from this submission.

