## Supplemental Materials for "Disease progression strikingly differs in research and real-world Parkinson’s populations"

### Supplemental Tables

#### Supplemental Table 1. Included PD medications.

Carbidopa (Lodosyn)

Artane (Trihexyphenidyl)

Carbidopa / Levodopa Immediate Release (Sinemet)

Benztropine (Cogentin)

Carbidopa / Levodopa Controlled Release (Sinemet CR)

Rotigotine Transdermal Patch (Neupro Patch)

Carbidopa / Levodopa Orally Disintegrating Tablets (Parcopa)

Northera (Droxidopa)

Carbidopa / Levodopa Extended Release Capsules (Rytary or Numient)

Nuplazid (Pimavanserin)

Carbidopa / Levodopa Intestinal Gel (Duopa or Duodopa)

Botox (Botulinum toxin)

Carbidopa / Levodopa and Entacapone (Stalevo)

Dysport (Botulinum toxin)

Entacapone (Comtan)

Myobloc (Botulinum toxin)

Rasagiline (Azilect)

Xeomin (Botulinum toxin)

Deprenyl (Selegiline)

Exelon (Rivastigmine)

Eldepryl (Selegiline)

Aricept (Donepezil)

Zelapar (Selegiline)

Reminyl (Galantamine)

Selegiline Transdermal (Emasm)

Razadyne (Galantamine)

Pramipexole (Mirapex)

Namenda or Namenda XR (Memantine)

Pramipexole Extended Release or Modified Release Tablets (Mirapex ER or Sifrol ER or Pramipexole XR GP)

Ebixa (Memantine)

Adartel (Ropinirole)

Requip (Ropinirole)

Ropinirole Extended Release (Requip XL)

Amantadine (Symmetrel)

Amantadine Extended Release (Gocovri ER or Osmolex ER)

Tolcapone (Tasmar)

Apomorphine (Apokyn)

Parlodel (Bromocriptine)

Safinamide (Xadago or Equfina)

Levodopa / Benserazide Immediate Release (Madopar or Prolopa)

Apo-Trihex (Trihexyphenidyl)

Apomorphine sublingual film (Kynmobi)

Carbidopa / Levodopa and Entacapone Intestinal Gel (Lecigon)

Carbidopa / Levodopa Inhalation Powder (Inbrija)

Levodopa / Benserazide Controlled Release (Madopar CR, Madopar HBS, or Prolopa CR)

Levodopa / Benserazide Dispersible (Madopar Rapid)

Ropinirole [Derived]

Ropinirole Transdermal Patch (Haruropi Tape or HP-3000)

Selegiline [Derived]

Trihexyphenidyl [Derived]

Ethopropazine (Parsitan or Parsidan or Profenamine or Parsidol, or Parkin)

Istradefylline (Nourianz or Nouriast)

Mucuna Pruriens

Opicapone (Ongentys)

#### Supplemental Table 2. Real World Data Definition of PD Diagnosis.

| Inclusion Codes | ICD9 | ICD10 |
| --- | --- | --- |
| Parkinson’s Disease | 332, 332.0 | G20 |
| Quiescence exclusion codes | ICD9 | ICD10 |
| Alzheimer’s Disease & Cerebral Degenerations | 331.* | G30* |
| Dementia | 290* | F03.90 |
| Multiple Systems Atrophy/Progressive Supranuclear Palsy | 333.0 | G90.3, G23.1 |
| Schizophrenia | 295* | F20* |
| Lewy Body Dementia | 331.82 | G31.83 |
| Encephalitis | 323* | G04* |
| Wilson’s Disease | 275.1 | E83.01 |

#### Supplemental Table 3. Included ICD codes for definition of each Clinical Event.

| Diagnosis |  | Codes |
| --- | --- | --- |
| Depression | ICD-9 | 296.2, 296.20, 296.21, 296.22, 296.23, 296.24, 296.25, 296.26,  296.3, 296.30, 296.31, 296.32, 296.33, 296.34, 296.35, 296.36,  296.82, 311 |
|  | ICD-10 | F32.9, F32.0, F32.1, F32.2, F32.3, F32.4, F32.5, F33.40,  F33.9, F33.0, F33.1, F33.2, F33.3, F33.40, F33.41, F33.42, F32.89 |
| Fractures | ICD-9 | 800.00, 800.01, 800.02, 800.03, 800.04, 800.05, 800.06, 800.09, 800.1, 800.11, 800.12, 800.13, 800.14, 800.15, 800.16, 800.19, 800.30, 800.31, 800.32, 800.34, 800.35, 800.36, 800.39, 800.4, 800.41, 800.42, 800.43, 800.45, 800.46, 800.49, 800.50, 800.51, 800.52, 800.53, 800.54, 800.55, 800.56, 800.59, 800.60, 800.61, 800.62, 800.63, 800.64, 800.65, 800.66, 800.69, 800.70, 800.71, 800.72, 800.73, 800.74, 800.75, 800.76, 800.79, 800.80, 800.81, 800.82, 800.83, 800.84, 800.85, 800.86, 800.89, 800.90, 800.91, 800.92, 800.93, 800.94, 800.95, 800.96, 800.99, 801.00, 801.01, 801.03, 801.04, 801.05, 801.06, 801.09, 801.10, 801.11, 801.12, 801.13, 801.14, 801.15, 801.16, 801.19, 801.20, 801.21, 801.22, 801.23, 801.24, 801.25, 801.26, 801.29, 801.30, 801.31, 801.32, 801.33, 801.34, 801.35, 801.36, 801.39, 801.40, 801.41, 801.42, 801.43, 801.44, 801.45, 801.46, 801.49, 801.50, 801.51, 801.52, 801.53, 801.54, 801.55, 801.56, 801.59, 801.60, 801.61, 801.62, 801.63, 801.64, 801.65, 801.66, 801.69, 801.70, 801.71, 801.72, 801.73, 801.74, 801.75, 801.76, 801.79, 801.80, 801.81, 801.82, 801.83, 801.84, 801.85, 801.86, 801.89, 801.90, 801.91, 801.92, 801.93, 801.94, 801.95, 801.96, 801.99, 802, 802.0, 802.1, 802.2, 802.20, 802.21, 802.22, 802.23, 802.24, 802.25, 802.26, 802.27, 802.28, 802.29, 802.3, 802.30, 802.31, 802.32, 802.33, 802.34, 802.35, 802.36, 802.37, 802.38, 802.39, 802.4, 802.5, 802.6, 802.7, 802.8, 802.9, 803.00, 803.01, 803.02, 803.04, 803.05, 803.06, 803.09, 803.10, 803.11, 803.12, 803.13, 803.14, 803.15, 803.16, 803.19, 803.20, 803.21, 803.22, 803.23, 803.24, 803.25, 803.26, 803.29, 803.30, 803.31, 803.32, 803.33, 803.34, 803.35, 803.36, 803.39, 803.40, 803.41, 803.42, 803.43, 803.44, 803.45, 803.46, 803.49, 803.50, 803.51, 803.52, 803.53, 803.54, 803.55, 803.56, 803.59, 803.60, 803.61, 803.62, 803.63, 803.64, 803.65, 803.66, 803.69, 803.70, 803.71, 803.72, 803.73, 803.74, 803.75, 803.76, 803.79, 803.80, 803.81, 803.82, 803.83, 803.84, 803.85, 803.86, 803.89, 803.90, 803.91, 803.92, 803.93, 803.94, 803.95, 803.96, 803.99, 804.00, 804.01, 804.02, 804.03, 804.04, 804.05, 804.06, 804.09, 804.10, 804.11, 804.12, 804.13, 804.14, 804.15, 804.16, 804.19, 804.20, 804.21, 804.22, 804.23, 804.24, 804.25, 804.26, 804.29, 804.30, 804.31, 804.32, 804.33, 804.34, 804.35, 804.36, 804.39, 804.40, 804.41, 804.42, 804.43, 804.44, 804.45, 804.46, 804.49, 804.50, 804.51, 804.52, 804.53, 804.54, 804.55, 804.56, 804.59, 804.60, 804.61, 804.62, 804.63, 804.64, 804.65, 804.66, 804.69, 804.70, 804.71, 804.72, 804.73, 804.74, 804.75, 804.76, 804.79, 804.80, 804.81, 804.82, 804.83, 804.84, 804.85, 804.86, 804.89, 804.90, 804.91, 804.92, 804.93, 804.94, 804.95, 804.96, 804.99, 805.00, 805.01, 805.02, 805.03, 805.04, 805.05, 805.06, 805.07, 805.08, 805.10, 805.11, 805.12, 805.13, 805.14, 805.15, 805.16, 805.17, 805.18, 805.2, 805.3, 805.4, 805.6, 805.7, 805.8, 805.9, 806, 806.00, 806.01, 806.02, 806.03, 806.04, 806.05, 806.06, 806.07, 806.08, 806.09, 806.1, 806.10, 806.11, 806.12, 806.13, 806.14, 806.15, 806.16, 806.17, 806.18, 806.19, 806.2, 806.20, 806.21, 806.22, 806.23, 806.24, 806.25, 806.26, 806.27, 806.28, 806.29, 806.3, 806.30, 806.31, 806.32, 806.33, 806.34, 806.35, 806.36, 806.37, 806.38, 806.39, 806.4, 806.5, 806.6, 806.60, 806.61, 806.62, 806.69, 806.7, 806.70, 806.71, 806.72, 806.79, 806.8, 806.9, 807, 807.00, 807.01, 807.02, 807.03, 807.04, 807.05, 807.06, 807.07, 807.08, 807.09, 807.10, 807.11, 807.12, 807.13, 807.14, 807.15, 807.16, 807.17, 807.18, 807.19, 807.2, 807.3, 808, 808.0, 808.1, 808.2, 808.3, 808.4, 808.43, 808.44, 808.49, 809, 809.0, 809.1, 810, 810.00, 810.01, 810.02, 810.03, 810.10, 810.11, 810.12, 810.13, 811.00, 811.01, 811.02, 811.03, 811.09, 811.10, 811.11, 811.12, 811.13, 811.19, 812, 812.00, 812.01, 812.02, 812.03, 812.10, 812.11, 812.12, 812.13, 813, 813.00, 813.01, 813.03, 813.04, 813.05, 813.06, 813.07, 813.08, 813.10, 813.11, 813.12, 813.13, 813.14, 813.15, 813.16, 813.17, 813.18, 813.20, 813.21, 813.22, 813.23, 813.30, 813.31, 813.32, 813.33, 813.40, 813.41, 813.42, 813.43, 813.44, 813.45, 813.46, 813.47, 813.50, 813.51, 813.52, 813.53, 813.54, 813.80, 813.81, 813.82, 813.83, 813.90, 813.91, 813.92, 813.93, 814.00, 814.01, 814.02, 814.03, 814.04, 814.05, 814.06, 814.07, 814.08, 814.09, 814.10, 814.11, 814.12, 814.13, 814.14, 814.15, 814.16, 814.17, 814.18, 814.19, 815.00, 815.01, 815.02, 815.03, 815.04, 815.09, 815.10, 815.11, 815.12, 815.13, 815.14, 815.19, 816.00, 816.01, 816.02, 816.03, 816.10, 816.11, 816.12, 816.13, 817.0, 817.1, 818.0, 818.1, 819.0, 819.1, 820.00, 820.01, 820.02, 820.03, 820.09, 820.10, 820.11, 820.12, 820.13, 820.19, 820.20, 820.21, 820.22, 820.30, 820.31, 820.32, 820.8, 820.9, 821.00, 821.01, 821.10, 821.11, 821.20, 821.21, 821.22, 821.23, 821.29, 821.30, 821.31, 821.32, 821.33, 821.39, 822, 822.1, 823.00, 823.01, 823.02, 823.10, 823.11, 823.12, 823.20, 823.21, 823.22, 823.30, 823.31, 823.32, 823.40, 823.41, 823.42, 823.80, 823.81, 823.82, 823.90, 823.91, 823.92, 824.0, 824.1, 824.2, 824.3, 824.4, 824.5, 824.6, 824.7, 824.8, 824.9, 825.0, 825.1, 825.20, 825.21, 825.22, 825.23, 825.24, 825.25, 825.29, 825.3, 825.31, 825.32, 825.33, 825.34, 825.35, 825.39, 826.0, 826.1, 827.0, 827.1, 828.0, 828.1, 829.0, 829.1 |
|  | ICD-10 | S02.0%%A, S02.0%%A, S06.9%1A, S06.9%2A, S02.0%%A, S06.9%3A, S06.9%4A, S02.0%%A, S06.9%5A, S02.0%%A, S06.9%6A, S06.9%9A, S06.0%9A, S02.0%%A, S06.319A, S06.339A, S06.330A, S02.0%%A, S06.330A, S02.0%%A, S06.331A, S06.332A, S06.313A, S06.314A, S06.323A, S06.324A, S06.333A, S06.334A, S02.0%%A, S06.335A, S02.0%%A, S06.336A, S06.337A, S06.338A, S06.339A, S06.319A, S06.329A, S06.330A, S06.339A, S02.0%%A, S02.0%%A, S06.309A, S06.360A, S02.0%%A, S06.300A, S06.360A, S06.301A, S06.302A, S06.361A, S06.362A, S06.305A, S06.365A, S06.306A, S06.366A, S06.367A, S06.368A, S02.0%%A, S06.309A, S06.369A, S02.0%%A, S06.300A, S06.309A, S06.360A, S02.0%%A, S06.890A, S06.9%0A, S02.0%%A, S06.890A, S06.9%0A, S02.0%%A, S06.891A, S06.892A, S06.9%1A, S06.9%2A, S02.0%%A, S06.893A, S06.894A, S06.9%3A, S06.9%4A, S02.0%%A, S06.896A, S06.897A, S06.898A, S06.9%6A, S06.9%7A, S06.9%8A, S02.0%%A, S06.899A, S06.9%9A, S02.0%%A, S06.899A, S06.9%9A, S02.0%%B, S06.9%1A, S02.0%%B, S02.0%%B, S02.0%%B, S02.0%%B, S06.9%5A, S02.0%%B, S02.0%%B, S02.0%%B, S02.0%%B, S02.0%%B, S02.0%%B, S02.0%%B, S02.0%%B, S02.0%%B, S02.0%%B, S02.0%%B, S02.0%%B, S02.0%%B, S02.0%%B, S02.0%%B, S02.0%%B, S02.0%%B, S02.0%%B, S02.0%%B, S02.0%%B, S02.0%%B, S02.0%%B, S02.0%%B, S02.0%%B, S02.0%%B, S02.0%%B, S02.0%%B, S02.0%%B, S02.0%%B, S02.0%%B, S02.0%%B, S02.0%%B, S02.0%%B, S02.0%%B, S02.0%%B, S02.101A, S02.102A, S02.109A, S02.110A, S02.111A, S02.112A, S02.113A, S02.118A, S02.119A, S02.11AA, S02.11BA, S02.11CA, S02.11DA, S02.11EA, S02.11FA, S02.11GA, S02.11HA, S02.19%A, S02.101A, S02.102A, S02.109A, S02.101A, S02.102A, S02.109A, S02.101A, S02.102A, S02.109A, S02.101A, S02.102A, S02.109A, S02.101A, S02.102A, S02.109A, S02.101A, S02.102A, S02.109A, S02.101A, S02.102A, S02.109A, S02.101A, S02.102A, S02.109A, S02.101A, S02.102A, S02.109A, S02.102B, S02.109B, S02.110B, S02.111B, S02.112B, S02.113B, S02.118B, S02.119B, S02.11AB, S02.11BB, S02.11CB, S02.11DB, S02.11EB, S02.11FB, S02.11GB, S02.11HB, S02.19%B, S02.101B, S02.101B, S02.102B, S02.109B, S02.101B, S02.102B, S02.109B, S02.101B, S02.102B, S02.109B, S02.101B, S02.102B, S02.109B, S02.101B, S02.102B, S02.109B, S02.2%%A, S02.2%%B, S02.609A, S02.69%A, S02.610A, S02.611A, S02.612A, S02.620A, S02.621A, S02.622A, S02.630A, S02.631A, S02.632A, S02.640A, S02.641A, S02.642A, S02.650A, S02.651A, S02.652A, S02.66%A, S02.670A, S02.671A, S02.672A, S02.600A, S02.601A, S02.602A, S02.609A, S02.69%A, S02.609B, S02.69%B, S02.611B, S02.612B, S02.610B, S02.620B, S02.621B, S02.622B, S02.630B, S02.631B, S02.632B, S02.640B, S02.641B, S02.642B, S02.650B, S02.651B, S02.652B, S02.66%B, S02.670B, S02.671B, S02.672B, S02.600B, S02.601B, S02.602B, S02.609B, S02.69%B, S02.401A, S02.402A, S02.40AA, S02.40BA, S02.40CA, S02.40DA, S02.40EA, S02.40FA, S02.411A, S02.412A, S02.413A, S02.400A, S02.401B, S02.402B, S02.40AB, S02.40BB, S02.40CB, S02.40DB, S02.40EB, S02.40FB, S02.411B, S02.412B, S02.413B, S02.400B, S02.30%A, S02.31%A, S02.32%A, S02.30%B, S02.31%B, S02.32%B, S02.42%A, S02.80%A, S02.81%A, S02.82%A, S02.92%A, S02.42%B, S02.80%B, S02.81%B, S02.82%B, S02.92%B, S02.91%A, S02.91%A, S02.91%A, S02.91%A, S02.91%A, S02.91%B, S02.91%B, S02.91%B, S02.91%B, S02.91%B, S02.91%A, S02.91%A, S02.91%A, S02.92%A, S02.91%A, S02.91%A, S02.91%B, S02.91%B, S02.91%B, S02.91%B, S02.91%B, S12.9%%A, S12.001A, S12.01%A, S12.02%A, S12.030A, S12.031A, S12.040A, S12.041A, S12.090A, S12.091A, S12.000A, S12.101A, S12.110A, S12.111A, S12.112A, S12.120A, S12.121A, S12.130A, S12.131A, S12.14%A, S12.150A, S12.151A, S12.190A, S12.191A, S12.100A, S12.201A, S12.230A, S12.231A, S12.24%A, S12.250A, S12.251A, S12.290A, S12.291A, S12.200A, S12.301A, S12.330A, S12.331A, S12.34%A, S12.350A, S12.351A, S12.390A, S12.391A, S12.300A, S12.401A, S12.430A, S12.431A, S12.44%A, S12.450A, S12.451A, S12.490A, S12.491A, S12.400A, S12.501A, S12.530A, S12.531A, S12.54%A, S12.550A, S12.551A, S12.590A, S12.591A, S12.500A, S12.601A, S12.630A, S12.631A, S12.64%A, S12.650A, S12.651A, S12.690A, S12.691A, S12.600A, S12.9%%A, S12.9%%A, S12.001B, S12.01%B, S12.02%B, S12.030B, S12.031B, S12.040B, S12.041B, S12.090B, S12.091B, S12.000B, S12.101B, S12.110B, S12.111B, S12.112B, S12.120B, S12.121B, S12.130B, S12.131B, S12.14%B, S12.150B, S12.151B, S12.190B, S12.191B, S12.201B, S12.230B, S12.231B, S12.24%B, S12.250B, S12.251B, S12.290B, S12.291B, S12.200B, S12.301B, S12.330B, S12.331B, S12.34%B, S12.350B, S12.351B, S12.390B, S12.391B, S12.300B, S12.401B, S12.430B, S12.431B, S12.44%B, S12.450B, S12.451B, S12.490B, S12.491B, S12.400B, S12.501B, S12.530B, S12.531B, S12.54%B, S12.550B, S12.551B, S12.590B, S12.591B, S12.500B, S12.601B, S12.630B, S12.631B, S12.64%B, S12.650B, S12.651B, S12.690B, S12.691B, S12.600B, S12.9%%A, S22.001A, S22.002A, S22.008A, S22.009A, S22.010A, S22.011A, S22.012A, S22.018A, S22.019A, S22.020A, S22.021A, S22.022A, S22.028A, S22.029A, S22.030A, S22.031A, S22.032A, S22.038A, S22.039A, S22.040A, S22.041A, S22.042A, S22.048A, S22.049A, S22.050A, S22.051A, S22.052A, S22.058A, S22.059A, S22.060A, S22.061A, S22.062A, S22.068A, S22.069A, S22.070A, S22.071A, S22.072A, S22.078A, S22.079A, S22.080A, S22.081A, S22.082A, S22.088A, S22.089A, S22.001B, S22.002B, S22.008B, S22.009B, S22.010B, S22.011B, S22.012B, S22.018B, S22.019B, S22.020B, S22.021B, S22.022B, S22.028B, S22.029B, S22.030B, S22.031B, S22.032B, S22.038B, S22.039B, S22.040B, S22.041B, S22.042B, S22.048B, S22.049B, S22.050B, S22.051B, S22.052B, S22.058B, S22.059B, S22.060B, S22.061B, S22.062B, S22.068B, S22.069B, S22.070B, S22.071B, S22.072B, S22.078B, S22.079B, S22.080B, S22.081B, S22.082B, S22.088B, S22.089B, S22.000B, S32.001A, S32.002A, S32.008A, S32.009A, S32.010A, S32.011A, S32.012A, S32.018A, S32.019A, S32.020A, S32.021A, S32.022A, S32.028A, S32.029A, S32.030A, S32.031A, S32.032A, S32.038A, S32.039A, S32.040A, S32.041A, S32.042A, S32.048A, S32.049A, S32.050A, S32.051A, S32.052A, S32.058A, S32.059A, S32.000A, S32.110A, S32.111A, S32.112A, S32.119A, S32.120A, S32.121A, S32.122A, S32.129A, S32.130A, S32.131A, S32.132A, S32.139A, S32.14%A, S32.15%A, S32.16%A, S32.17%A, S32.19%A, S32.2%%A, S32.10%A, S32.110B, S32.111B, S32.112B, S32.119B, S32.120B, S32.121B, S32.122B, S32.129B, S32.130B, S32.131B, S32.132B, S32.139B, S32.14%B, S32.15%B, S32.16%B, S32.17%B, S32.19%B, S32.2%%B, S32.10%B, S12.9%%A, S22.009A, S32.009A, S32.10%A, S32.2%%A, S22.009B, S32.009B, S32.10%B, S32.2%%B, S12.001A, S12.100A, S12.101A, S12.200A, S12.201A, S12.300A, S12.301A, S12.9%%A, S12.000A, S12.401A, S12.500A, S12.501A, S12.600A, S12.601A, S12.400A, S12.001B, S12.100B, S12.101B, S12.200B, S12.201B, S12.300B, S12.301B, S12.9%%A, S12.000B, S12.401B, S12.500B, S12.501B, S12.600B, S12.601B, S12.400B, S22.019A, S22.029A, S22.039A, S22.049A, S22.059A, S22.009A, S22.069A, S22.079A, S22.089A, S22.009A, S22.019B, S22.029B, S22.039B, S22.049B, S22.059B, S22.009B, S22.069B, S22.079B, S22.089B, S22.009B, S32.019A, S32.029A, S32.039A, S32.049A, S32.059A, S32.009A, S32.2%%A, S32.10%A, S32.2%%B, S32.10%B, S22.009A, S12.9%%A, S32.009A, S32.10%A, S12.9%%A, S22.009B, S32.009B, S32.10%B, S22.39%A, S22.31%A, S22.32%A, S22.39%A, S22.42%A, S22.43%A, S22.49%A, S22.41%A, S22.39%B, S22.32%B, S22.39%B, S22.31%B, S22.42%B, S22.43%B, S22.49%B, S22.41%B, S22.21%A, S22.22%A, S22.23%A, S22.24%A, S22.20%A, S22.21%B, S22.22%B, S22.23%B, S22.24%B, S22.20%B, S32.402A, S32.409A, S32.411A, S32.412A, S32.413A, S32.414A, S32.415A, S32.416A, S32.421A, S32.422A, S32.423A, S32.424A, S32.425A, S32.426A, S32.431A, S32.432A, S32.433A, S32.434A, S32.435A, S32.436A, S32.441A, S32.442A, S32.443A, S32.444A, S32.445A, S32.446A, S32.451A, S32.452A, S32.453A, S32.454A, S32.455A, S32.456A, S32.461A, S32.462A, S32.463A, S32.464A, S32.465A, S32.466A, S32.471A, S32.472A, S32.473A, S32.474A, S32.475A, S32.476A, S32.481A, S32.482A, S32.483A, S32.484A, S32.485A, S32.402B, S32.409B, S32.411B, S32.412B, S32.413B, S32.414B, S32.415B, S32.416B, S32.421B, S32.422B, S32.423B, S32.424B, S32.425B, S32.426B, S32.431B, S32.432B, S32.433B, S32.434B, S32.435B, S32.436B, S32.441B, S32.442B, S32.443B, S32.444B, S32.445B, S32.446B, S32.451B, S32.452B, S32.453B, S32.454B, S32.455B, S32.456B, S32.461B, S32.462B, S32.463B, S32.464B, S32.465B, S32.466B, S32.471B, S32.472B, S32.473B, S32.474B, S32.475B, S32.476B, S32.481B, S32.482B, S32.483B, S32.484B, S32.485B, S32.502A, S32.509A, S32.511A, S32.512A, S32.519A, S32.591A, S32.592A, S32.599A, S32.501A, S32.502B, S32.509B, S32.511B, S32.512B, S32.519B, S32.591B, S32.592B, S32.599B, S32.501B, S32.810A, S32.811A, S32.82%A, S32.89%A, S32.9%%A, S22.9%%A, S22.9%%B, S42.002A, S42.009A, S42.001A, S42.012A, S42.013A, S42.014A, S42.015A, S42.016A, S42.017A, S42.018A, S42.019A, S42.022A, S42.023A, S42.024A, S42.025A, S42.026A, S42.021A, S42.032A, S42.033A, S42.034A, S42.035A, S42.036A, S42.031A, S42.002B, S42.009B, S42.001B, S42.012B, S42.013B, S42.014B, S42.015B, S42.016B, S42.017B, S42.018B, S42.019B, S42.022B, S42.023B, S42.024B, S42.025B, S42.026B, S42.032B, S42.033B, S42.034B, S42.035B, S42.036B, S42.031B, S42.101A, S42.102A, S42.109A, S42.122A, S42.123A, S42.124A, S42.125A, S42.126A, S42.121A, S42.131A, S42.132A, S42.133A, S42.134A, S42.135A, S42.136A, S42.142A, S42.143A, S42.144A, S42.145A, S42.146A, S42.151A, S42.152A, S42.153A, S42.154A, S42.155A, S42.156A, S42.112A, S42.113A, S42.114A, S42.115A, S42.116A, S42.191A, S42.192A, S42.199A, S42.101B, S42.102B, S42.109B, S42.122B, S42.123B, S42.124B, S42.125B, S42.126B, S42.121B, S42.132B, S42.133B, S42.134B, S42.135B, S42.136B, S42.142B, S42.143B S42.144B, S42.145B, S42.146B, S42.151B, S42.152B, S42.153B, S42.154B, S42.155B, S42.156B, S42.141B, S42.112B S42.113B, S42.114B, S42.115B, S42.116B, S42.191B, S42.192B, S42.199B, S42.111B, S42.201A, S42.202A, S42.209A S42.212A, S42.213A, S42.214A, S42.215A, S42.216A, S42.221A, S42.222A, S42.223A, S42.224A, S42.225A, S42.226A, S42.231A, S42.232A, S42.239A, S42.241A, S42.242A, S42.249A, S42.211A, S42.291A, S42.292A, S42.293A, S42.294A, S42.295A, S42.296A, S42.252A, S42.253A, S42.254A, S42.255A, S42.256A, S42.202B, S42.209B, S42.201B, S42.212B, S42.213B, S42.214B, S42.215B, S42.216B, S42.221B, S42.222B, S42.223B, S42.224B, S42.225B, S42.226B, S42.231B, S42.232B, S42.239B, S42.241B, S42.242B, S42.249B, S42.211B, S42.292B, S42.293B, S42.294B, S42.295B, S42.296B, S42.291B, S42.252B, S42.253B, S42.254B, S42.255B, S42.256B, S42.251B, S52.002A, S52.009A, S52.101A, S52.102A, S52.109A, S52.90%A, S52.001A, S52.022A, S52.023A, S52.024A, S52.025A, S52.026A, S52.031A, S52.032A, S52.033A, S52.034A, S52.035A, S52.036A, S52.021A, S52.271A, S52.272A, S52.279A, S52.002A, S52.009A, S52.091A, S52.092A, S52.099A, S52.001A, S52.121A, S52.122A, S52.123A, S52.124A, S52.125A, S52.126A, S52.133A, S52.136A, S52109A, S52.189A, S52.009A, S52.109A, S52.90%B, S52.90%C, S52.023B, S52.023C, S52.026B, S52.026C, S52043B, S52.043C, S52.046B, S52.046C, S52.279B, S52.009B, S52.009C, S52.099B, S52.099C, S52.123B, S52123C, S52.126B, S52.126C, S52.133B, S52.133C, S52.136B, S52.136C, S52.109B, S52.109C, S52.189B, S52.189C, S52.009B, S52.009C, S52.109B, S52.109C, S52.90%A, S52.309A, S52.209A, S52.209A, S52.309A, S52.90%B, S5290%C, S52.309B, S52.309C, S52.209B, S52.209C, S52.209B, S52.209C, S52.309B, S52.309C, S52.90%A, S52539A, S52.549A, S52.509A, S52.609A, S52.509A, S52.119A, S52.529A, S52.019A, S52.629A, S52.521A, S52.621A, S52.011A, S52.111A, S52.522A, S52.622A, S52.012A, S52.112A, S52.90%B, S52.90%C, S52.539B, S52.539C, S52.509B, S52.509C, S52.609B, S52.609C, S52.509B, S52.509C, S52.609B, S52.609C, S52.90%A, S52.90%A, S52.90%A, S52.90%A, S52.90%B, S52.90%C, S62.109A, S62.009A, S62.123A, S62.126A, S62.113A, S62.116A, S62.163A, S62.166A, S62.173A, S62.176A, S62.183A, S62.186A, S62.133A, S62.136A, S62.143A, S62.146A, S62.153A, S62.156A, S62.109B, 62.009B, S62.123B, S62.126B, S62.113B, S62.116B, S62.163B, S62.166B, S62.173B, S62.176B, S62.183B, S62.186B, S62.133B, S62.136B, S62.143B, S62.146B, S62.153B, S62.156B, S62.309A, S62.233A, S62.236A, S62.319A, S62.349A, S62.329A, 62.359A N, S62.339A, S62.369A, S62.399A, S62.309B, S62.233B, S62.236B, S62.319B, S62.349B, S62.329B, S62.359B, S62.339B, S62.369B, S62.399B, S62.509A, S62.609A, S62.513A, S62.516A, S62.629A, S62.649A, S62.659A, S62.523A, S62.526A, S62.639A, S62.669A, S62.90%A, S62.509B, S62.609B, S62.513B, S62.516B, S62.619B, S62.629B, S62.649B, S62.659B, S62.523B, S62.526B, S62.639B, S62.669B, S62.90%B, S62.90%A, S62.90%B, S62.90%A, S62.90%B, S42.91%A, S52.91%A, S42.92%A, S52.92%A, S22.20%A, S22.49%A, S42.90%A, S52.90%A, S42.91%B, S52.91%B, S42.92%B, S52.92%B, S22.20%B, S22.49%B, S42.90%B, S52.90%B, S72.019A, S72.023A, S72.026A, S72.033A, S72.036A, S72.043A, S72.046A, S72.099A, S72.019B, S72.019C, S72.023B, S72.023C, S72.026B, S72.026C, S72.033B, S72.033C, S72.036B, S72.036C, S72.043B, S72.043C, S72.046B, S72.046C, S72.099B, S72.099C, S72.109A, S72.143A, S72.146A, S72.23%A, S72.26%A, S72.109B, S72.109C, S72.143B, S72.143C, S72.146B, S72.146C, S72.23%B, S72.23%C, S72.26%B, S72.26%C, S72.009A, S72.009B, S72.009C, S72.90%A, S72.309A, S72.90%B, S72.90%C, S72.309B, S72.309C, S72.409A, S72.413A, S72.416A, S72.443A, S72.446A, S72.453A, S72.456A, S72.499A, S72.409B, S72.409C, S72.413B, S72.413C, S72.416, S72.416C, S72.443B, S72.443C, S72.446, S72.446C, S72.453B, S72.453C, S72.456B, S72.456C, S72.499B, S72.499C, S82.009A, S82.009B, S82.009C, S82.109A, S82.839A, S82.101A, S82.831A, S82.102A, S82.832A, S82.109B, S82.109C, S82.839B, S82.839C, S82.101B, S82.831B, S82.102B, S82.832B, S82.209A, S82.409A, S82.201A, S82.401A, S82.202A, S82.402A, S82.209B, S82.209C, S82.409B, S82.409C, S82.201B, S82.401B, S82.202B, S82.169A, S82.819A, S82.161A, S82.811A, S82.311A, S82.821A, 162A Toru, S82.812A, S82.312A, S82.822A, S82.201A, S82.401A, S82.201A, S82.401A, S82.201B, S82.401C, S82.401B, S82.401C, S82.53%A, S82.56%A, S82.53%B, S82.53%C, S82.56%B, S82.56%C, S82.63%A, S82.66%A, S82.63%B, S82.63%C, S82.66%B, S82.66%C, S82.843, S82.843B, S82.843C, S82.846B, S82.846C, S82.853A, S82.856A, S82.846B, S82.846C, S82.899A, S82.899B, S82.899C, S92.009A, S99.009A, S99.019A, S99.029A, S99.039A, S99.049A, S99.099A, S92.009B, S99.009B, S99.019B, S99.029B, S99.039B, S99.049B, S99.099B, S92.819A, S92.909A, S92.109A, S92.253A, S92.256A, S92.213A, S92.223A, S92.226A, S92.309A, S92.201A, S92.202A, S92.209A, S92.819B, S92.109B, S92.253B, S92.256B, S92.213B, S92.216B, S92.223B, S92.226B, S92.309B, S92.201B, S92.202B, S92.209B, S92.403A, S92.406A, S92.503A, S92.506A, S92.403B, S92.406B, S92.503B, S92.506B, S82.90%A, S82.90%B, S72.91%A, S82.91%A, S72.92%A, S82.92%A, S42.90%A, S52.90%A, S72.90%A, S82.90%A, S22.20%A, S22.49%A, S72.90%A, S82.90%A, S72.91%E, S82.91%B, S72.92%E, S82.92%B, S42.90%B, S52.90%B, S72.90%E, S82.90%B, S22.20%B, S22.49%B, S72.90%E, S82.90%B, T14.8%%A, T14.8%%A |
| Falls | ICD-9 | E881.1, E882, E883.0, E883.1, E883.2, E883.9, E884, E884.0, E884.1, E884.2, E884.3, E884.4, E884.5, E884.6, E884.9, E885.9, E886, E886.0, E886.9, E888, E888.0, E888.1, E888.8, E888.9 |
|  | ICD-10 | W10.0%%A, W10.0%%D, W10.0%%S, W10.1%%A, W10.1%%D, W10.1%%S, W10.2%%A, W10.2%%D, W10.8%%A, W10.8%%D, W10.8%%S, W10.9%%A, W10.9%%D, W11.%%%A, W11.%%%D, W11.%%%S, W12.%%%A, W12.%%%D, W12.%%%S, W13.0%%A, W13.0%%D, W13.1%%A, W13.1%%D, W13.2%%A, W13.2%%D, W13.3%%A, W13.3%%D, W13.4%%A, W13.4%%D, W13.8%%A, W13.8%%D, W13.9%%A, W13.9%%D, W16.011A, W16.011D, W16.012A, W16.012D, W16.021A, W16.021D, W16.022A, W16.022D, W16.031A, W16.031D, W16.032A, W16.032D, W16.111A, W16.111D, W16.112A, W16.112D, W16.121A, W16.121D, W16.122A, W16.122D, W16.131A, W16.131D, W16.132A, W16.132D, W16.211A, W16.211D, W16.212A, W16.212D, W16.221A, W16.221D, W16.222A, W16.222D, W16.311A, W16.311D, W16.312A, W16.312D, W16.321A, W16.321D, W16.322A, W16.322D, W16.331A, W16.331D, W16.332A, W16.332D, W16.41%A, W16.41%D, W16.42%A, W16.42%D, W16.511A, W16.511D, W17.0%%A, W17.0%%D, W17.0%%S, W17.1%%A, W17.1%%D, W17.1%%S, W17.2%%A, W17.2%%D, W17.3%%A, W17.3%%D, W17.4%%A, W17.4%%D, W09.0%%A, W09.0%%D, W09.1%%A, W09.1%%D, W09.2%%A, W09.2%%D, W09.8%%A, W09.8%%D, W15.%%%A, W15.%%%D, W15.%%%S, W07.%%%A, W07.%%%D, W07.%%%S, V00.811A, V00.811D, V00.812A, V00.812D, V00.818A, V00.818D, V00.831A, V00.831D, V00.832A, V00.832D, V00.838A, V00.838D, W05.0%%A, W05.0%%D, W05.1%%A, W05.1%%D, W05.2%%A, W05.2%%D, W06.%%%A, W06.%%%D, W06.%%%S, V00.821A, V00.821D, V00.822A, V00.822D, V00.828A, V00.828D, W08.%%%A, W08.%%%D, W08.%%%S, W18.11%A, W18.11%D, W18.12%A, W18.12%D, V00.891A, V00.891D, V00.892A, V00.892D, V00.898A, V00.898D, W00.1%%A, W00.1%%D, W00.2%%A, W00.2%%D, W14.%%%A, W14.%%%D, W17.81%A, W17.81%D, W17.82%A, W17.82%D, W17.89%A, W17.89%D, V00.181A, V00.181D, V00.182A, V00.182D, V00.211A, V00.211D, V00.212A, V00.212D, V00.221A, V00.221D, V00.222A, V00.222D, V00.281A, V00.281D, V00.282A, V00.282D, V00.381A, V00.381D, V00.382A, V00.382D, W00.0%%A, W00.0%%D, W00.9%%A, W00.9%%D, W01.0%%A, W01.0%%D, W18.2%%A, W18.2%%D, W18.40%A, W18.40%D, W18.41%A, W18.41%D, W18.42%A, W18.42%D, W18.43%A, W18.43%D, W18.49%A, W18.49%D, W03.%%%A, W03.%%%D, V00.188A, V00.188D, V00.218A, V00.218D, V00.228A, V00.228D, V00.288A, V00.288D, V00.388A, V00.388D, W01.10%A, W01.10%D, W01.110A, W01.110D, W01.111A, W01.111D, W01.118A, W01.118D, W01.119A, W01.119D, W18.02%A, W18.02%D, W01.190A, W01.190D, W01.198A, W01.198D, W18.00%A, W18.00%D, W18.01%A, W18.01%D, W18.09%D, W04.%%%A, W04.%%%D, W18.30%A, W18.30%D, W18.31%A, W18.31%D, W18.39%A, W18.39%D, W18.39%S, W19.%%%A, W19.%%%D, W19.%%%S, V00.181A, V00.181D, V00.182A, V00.182D, V00.211A, V00.211D, V00.212A, V00.212D, V00.221A, V00.221D, V00.222A, V00.222D, V00.281A, V00.281D, V00.282A, V00.282D, V00.381A, V00.381D, V00.382A, V00.382D, W00.0%%A, W00.0%%D, W00.9%%A, W00.9%%D, W01.0%%A, W01.0%%D, W18.2%%A, W18.2%%D, W18.40%A, W18.40%D, W18.41%A, W18.41%D, W18.42%A, W18.42%D, W18.43%A, W18.43%D, W18.49%A, W18.49%D |

#### Supplemental Table 4. Regression of Clinical Rating Scale by number of years from PD onset.

|  | **Data Source** | **Slope** | **Intercept** | **r^2^ Coefficient** | **p-value for # years** | **p-value between groups** |
| --- | --- | --- | --- | --- | --- | --- |
| **H&Y** | **HBS** | 0.05 | 1.99 | 0.30 | < 0.001 | > 0.05 |
|  | **MGB** | 0.07 | 2.18 | 0.24 | < 0.001 |  |
| **MMSE** | **HBS** | -0.11 | 28.67 | -0.17 | < 0.001 | < 0.001 |
|  | **MGB** | -0.28 | 25.81 | -0.14 | <0.001 |  |
| **UPDRS Total** | **HBS** | 1.54 | 28.25 | 0.37 | < 0.001 | < 0.001 |
|  | **MGB** | 3.87 | 18.93 | 0.29 | < 0.001 |  |

#### Supplemental Table 5. Comparison of Hoehn & Yahr progression.

| Hoehn & Yahr Stage | MGB Median Months to Transition to Next Stage | Zhao et al.^1^ Median Months to Transition to Next Stage |
| --- | --- | --- |
| 1 | 18 | 20 |
| 2 | 57 | 62 |
| 2.5 | 29 | 25 |
| 3 | 20 | 24 |
| 4 | 27 | 26 |
| 5 | NA | NA |
| Weighted Expected annual scale increase per year: | **0.39** | **0.45** |

#### Supplemental Table 6. Percentage of Patients with Levodopa Initiation By H&Y Stage (HBS).

| **H&Y Level** | **Levodopa Init** | **Total at HY** | **% Levodopa Init** |
| --- | --- | --- | --- |
| **1** | 69 | 136 | 50.7% |
| **1.5** | 47 | 74 | 63.5% |
| **2** | 782 | 1141 | 68.5% |
| **2.5** | 373 | 458 | 81.4% |
| **3** | 210 | 261 | 80.5% |
| **4** | 48 | 63 | 76.2% |
| **5** | 19 | 23 | 82.6% |

### Supplemental Figures

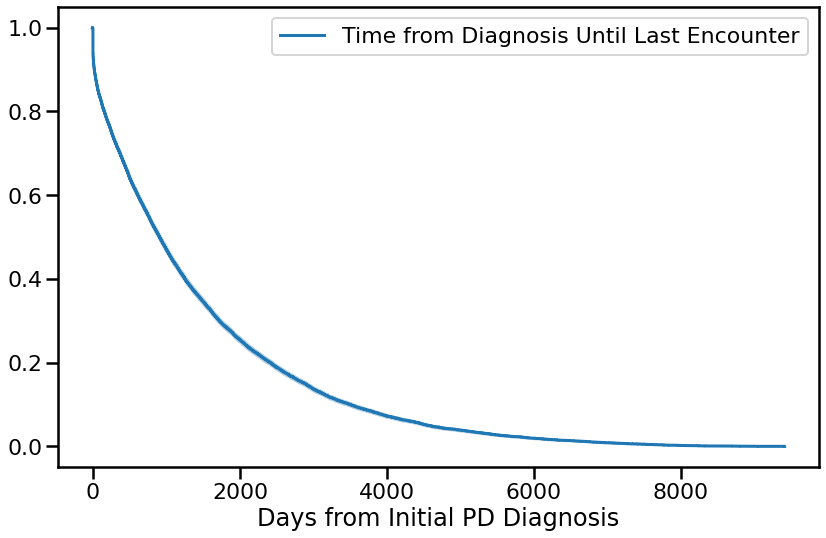

#### Supplemental Figure 1. MGB Time from initial PD Diagnosis until last encounter.
